# Effectiveness of ten-valent pneumococcal conjugate vaccine on invasive pneumococcal disease among children <2 years old: a prospective population-based study in rural Bangladesh

**DOI:** 10.1101/2023.07.23.23292706

**Authors:** Roly Malaker, Md Hassanuzaman, Hafizur Rahman, Rajib Chandra Das, Naito Kanon, Senjuti Saha, Yogesh Hooda, Arif M Tanmoy, Sowmitra Ranjan Chakraborty, Shampa Saha, Maksuda Islam, Gary L. Darmstadt, Abdullah H Baqui, Mathuram Sathosam, Shams El-Arifeen, Cynthia G Whitney, Samir K Saha

## Abstract

The 10-valent pneumococcal conjugate vaccine (PCV10) was introduced in March 2015 in Bangladesh. In this study, we aimed to estimate the impact of PCV10 on invasive pneumococcal disease (IPD) identified by blood cultures and severe pneumonia identified clinically and its effectiveness on invasive disease caused by vaccine serotypes.

We conducted population-based surveillance among children aged 2- <24 months between April 2012 through March 2019 in Mirzapur, a rural sub-district of Bangladesh. We compared incidence of IPD and severe pneumonia before (April 2012 to March 2015) and after (April 2015 to March 2019) the introduction of PCV10. Vaccine effectiveness was measured using an indirect cohort analysis of data from four sentinel sites in which PCV10 vaccination status was compared among children with IPD caused by vaccine serotype vs. non-vaccine serotypes.

We identified 24 IPD cases by blood culture and 1,704 severe pneumonia hospitalizations during the surveillance period. IPD incidence in under-2-year-old children fell 25% (95% CI: -1.2% to 76%; p-value =0.59) from 106 cases per 100,000 person-years at baseline to 79.3 in April 2018- March 2019. Vaccine serotype-IPD incidence was lower (77% reduction, 95% CI: -0.45% to 96%; p-value =0.068) in April 2018 - March 2019 than in the pre-vaccine period (85.7 cases to 19.8/100,000 observation-years). A significant decline of 54.0% (95% CI: 47.0% to 59.0%; p-value <0.001) was observed in hospitalizations due to severe pneumonia. From indirect cohort analysis, the effectiveness of PCV10 against vaccine serotype IPD was 37% (95% CI: -141.0% to 83.5%; p= 0.5) after the 1^st^ dose and 63.1% (95% CI: -3.3% to 85.9%, p=0.0411) after the 2^nd^ or the 3^rd^ dose.

This study demonstrates that PCV10 introduction prevented hospitalizations with severe pneumonia and provided individual protection against vaccine serotypes.

## Introduction

*Streptococcus pneumoniae* is the leading cause of childhood invasive diseases, including pneumonia, sepsis, and meningitis, worldwide (1, 2). Approximately 45 million episodes of severe pneumococcal disease occurred in 2016, leading to 350,000 deaths among children aged <5 years (3, 4). According to an estimate from 2000, 21,000 deaths occur annually in Bangladesh from pneumococcal disease (2). Effective vaccines against *S. pneumoniae* became available for children in 2000, and the World Health Organization (WHO) recommended the inclusion of Pneumococcal Conjugate Vaccine (PCV) in childhood immunization programs in 2006 (5). However, lack of data on the impact of immunization in low-income settings and the limited number of serotypes included in the first formulation (PCV7) hindered institution of data-driven decisions by policymakers from Gavi-eligible countries, which suffer the highest disease burden (2, 3). PCV was initially introduced in the USA and other affluent countries where it led to significant reductions in invasive pneumococcal disease (IPD) among children (3, 6-8). Many countries, however, observed the emergence of non-vaccine serotypes (NVTs), which partially offset the success of vaccination (9). Because of this observation, WHO emphasized the importance of monitoring the impact of PCVs after their introduction into routine vaccination programs (10).

We have been generating data from both hospital- and population-based surveillance since 2004 in Bangladesh. These data have been used to estimate the burden of pneumococcal disease in South Asia (11-13). Since 2004, our team has maintained both community- and hospital-based surveillance in Mirzapur, a rural region in Bangladesh, to assess the burden of IPD (13), in addition to conducting hospital-based surveillance in three additional sites across the country. The Bangladesh government introduced PCV10 (Synflorix, GSK) into its Expanded Program on Immunization (EPI) in March 2015, to children in a 3-dose primary series at 6, 10 and 14 weeks after birth.

In this study, we measured vaccine impact on pneumococcal disease in children aged <2 years in Mirzapur subdistrict by comparing data from the pre-vaccine period (April 2012 - March 2015) to that from the post-vaccine period (April 2015-March 2019). Pneumococcal pneumonia is mostly non-bacteremic in this setting, and thus isolation of this organism in the bloodstream is rare. Therefore, we aimed: 1) to measure the impact of vaccine by comparing incidence before and after introduction of PCV10 on: a) the number of laboratory-confirmed IPD cases, and b) the number of severe pneumonia cases identified clinically by physicians among children <2 years of age, and 2) to analyze the effectiveness of PCV10 against vaccine type (VT)-IPD using an indirect cohort method and data from laboratory-based surveillance.

## Methods

### Study design

To achieve the study objectives, we used two different study design approaches: (1) a before-after study in Mirzapur based on community surveillance for suspected pneumococcal disease and hospital case detection of blood culture-proven IPD at the population level to compare: a) overall, vaccine type (VT) and non-vaccine type (NVT) IPD incidence, and b) incidence of severe pneumonia, and (2) an indirect cohort study design using four sites, including the Mirzapur site, to assess the effectiveness of the PCV-10 against VTs, with IPD cases identified in Mirzapur and three other surveillances sites of the World Health Organization (WHO)-supported Invasive Bacterial Vaccine-Preventable Disease Surveillance Network sentinel site hospitals (11).

### Study site, population, and surveillance system

Population-based surveillance was conducted at Mirzapur, Bangladesh, 65 km north of Dhaka. In 2011, Mirzapur had an estimated total population of 400,000, distributed in 14 unions and 219 villages. The Kumudini Women’s Medical College Hospital (KWMCH) is the main health care facility for the population of Mirzapur. The study hospital (KWMCH) is the largest (1280-bed) non-profit private hospital in Bangladesh.

Ten of the 14 unions (Figure 1A) were covered by the demographic surveillance system. This demographic surveillance population is an open cohort; information on vital events like migration, pregnancy outcomes, births and deaths were recorded every fourth month. Children residing within the study area were eligible for this community-based surveillance. Trained Village Health Workers (VHWs) enrolled children in the community surveillance soon after birth once the primary guardian consented to the child’s participation. At enrollment, each baby was assigned a unique barcode which was printed on a smart card and remained valid until the child turned 5 years of age. The barcoded smart cards were used to identify the children and their study area. Most of the <5 children received care at the study hospital (Figure 1B), which was provided free of charge through Child Health Research Foundation (CHRF) support.

**Figure 1:**
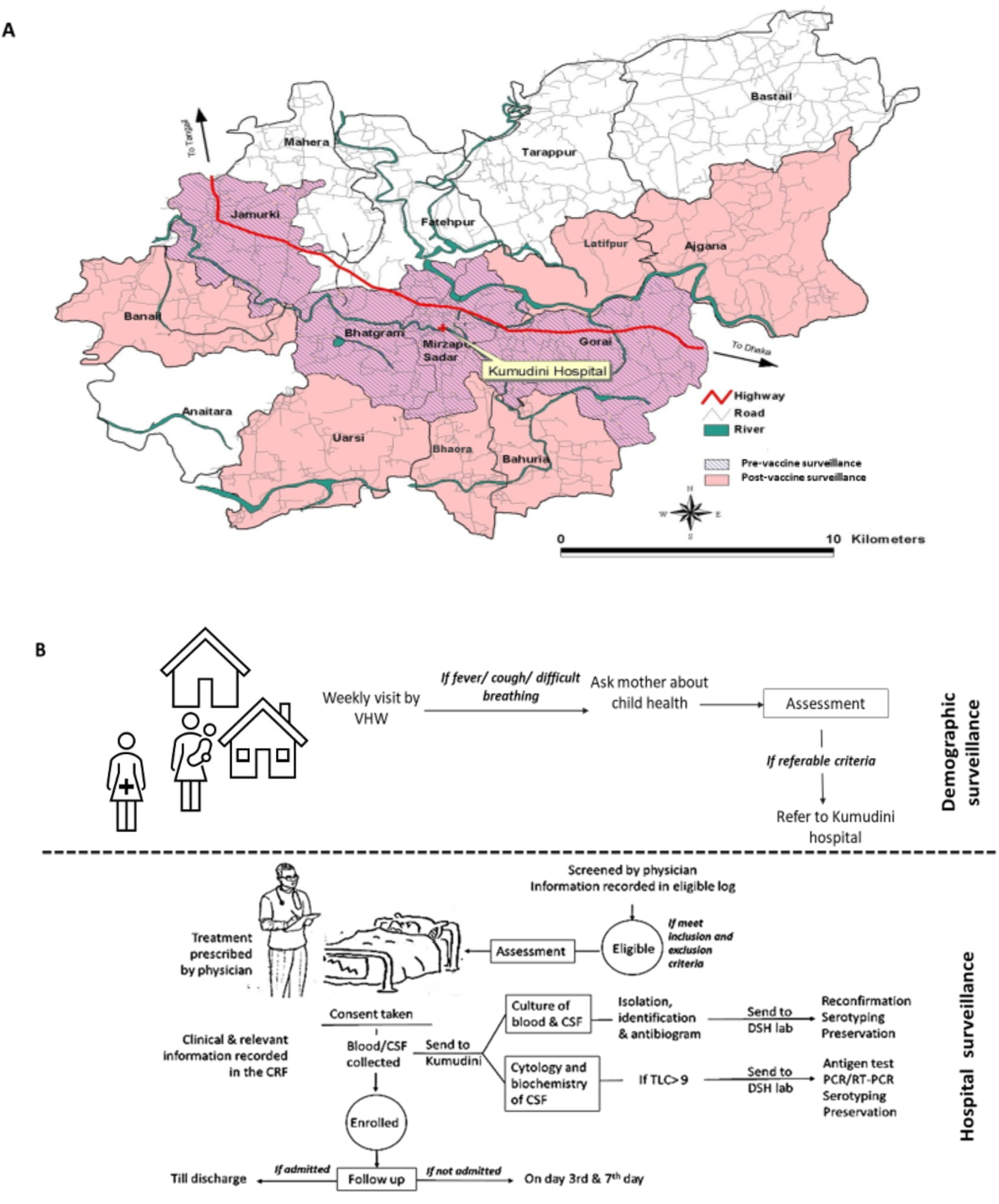
A. Demographic surveillance area of Mirzapur Upazila, Tangail; B. Study outline of community and hospital-based surveillance.

In the pre-vaccine period, surveillance was carried out in four unions; during the post-vaccine period, surveillance was extended to six additional unions, for a total of ten (Figure 1A). VHWs visited all children in their designated area once a week and inquired about the child’s well-being. If the mother/caregiver reported that a child had fever, cough or difficulty breathing, the VHW assessed the sick child using an algorithm (Table 1). The child was referred to KWMCH (Figure 1B) for medical care if the child met criteria provided in the algorithm for signs and symptoms of possible severe pneumonia/very severe disease, or high fever/possible sepsis. Children who were referred by VHWs or brought to the clinic due to family concerns of illness were evaluated clinically by a study physician who provided appropriate care and obtained specimens as indicated for identification of IPD.

**Table 1:** Case definition and management of children after assessment by VHWs in the Mirzapur community.

| Classification | Sign/ Symptoms | Management |
| --- | --- | --- |
| Possible severe Pneumonia/ Very Severe Disease | Cough, difficult breathing and/or fever (<99.5°F), with one of the following:<br>Convulsion<br>Fast breathing<br>Chest indrawing<br>Abnormally sleepy or difficult to wake<br>Unconscious<br>Not able to feed breast milk or drink at all<br>Vomits everything | Urgent refer to Kumudini hospital |
| Possible Pneumonia | Fast breathing, but no other danger sign | Refer to nearby health facilities |
| No pneumonia | No fast breathing, no danger sign | Home care |
| Low Fever | Temperature 99.5°F - 100.3°F | Home care |
| Moderate Fever | Temperature 100.4°F - 101.9°F | Urgent refer to Kumudini hospital |
| High fever/ Possible bacteremia | Temperature 102°F or above | Urgent refer to Kumudini hospital |
| Suspected Meningitis / Very severe disease | Fever (temperature 99.5°F and above) and at least one of the following:<br>▪ History of Stiff neck (mentioned by mother)<br>▪ Bulging fontanelle (if < 12 months of age)<br>▪ Unable to drink or breast feed<br>▪ Convulsions<br>▪ Lethargy (abnormally sleepy or difficult to wake)<br>▪ Unconscious<br>▪ Vomits everything | Urgent refer to Kumudini hospital |

**Table 2:** PCV10 effectiveness estimation by number of doses among children 2- <24 months using the indirect cohort method.

|  | Doses | Number of children | VT case/Total (VT-vaccinated+ unvaccinated) | NVT case/Total (NVT-vaccinated+ NVT-unvaccinated) | Crude vaccine effectiveness | p-value |
| --- | --- | --- | --- | --- | --- | --- |
| Overall | Unvaccinated children | 22 | 7 | 15 | n/a | n/a |
|  | 1st dose | 22 | 5/12 (41.67%) | 17/32 (53.13%) | 37.0% (-141.0%, 83.5%) | 0.498 |
|  | 2nd dose + 3rd dose | 75 | 9/21 (42.86%) | 65/97 (67.01%) | 63.0% (3.3%, 85.9%) | 0.041 |
|  | Any dose | 96 | 14/21 (66.67%) | 82/97 (84.54%) | 63.4% (-5.7%, 87.34%) | 0.072 |

Specimens (blood and/or CSF) for identification of IPD were collected from children who met the case definition of pneumonia, severe pneumonia, meningitis, and/or sepsis according to the WHO. The following case definitions were used by the study physicians of all four hospitals in the surveillance network to identify children with suspected pneumococcal disease in whom blood cultures were obtained for confirmation of IPD:

a. *Pneumonia:* Pneumonia was defined as history of cough or breathing difficulty with fast breathing (respiration rate ≥50/min for children 2- <12 months of age, ≥ 40/min for children ≥12 months of age).
b. *Severe pneumonia (physician confirmed):* A case of severe pneumonia was defined as history of cough or breathing difficulty and presence of any of the following danger signs (chest indrawing, inability to drink/breast feed, substantial vomiting, convulsions, lethargy, or fast breathing (respiratory rate ≥ 60 breaths/min in children <2 months of age), stridor in a calm child, altered consciousness).
c. *Meningitis:* A case of meningitis was defined as sudden onset of fever (>100.4°F) and presence of at least one of the following: neck stiffness, altered consciousness, bulging fontanel if <12 months of age, lethargy, convulsions, toxic appearance, petechial/purpural rash, poor sucking, or irritability.
d. *Sepsis:* A case of sepsis was defined as presence of any of the following danger signs (inability to drink/breast feed, convulsions, lethargy, stridor in a calm child, persistent vomiting, or severe malnutrition) excluding pneumonia and meningitis features.

Children aged 2 – <24 months who presented to the four study hospitals with suspected pneumococcal disease based on clinical examination were enrolled in the hospital surveillance portion of the study if their parents or legal guardian provided informed written consent for recording clinical features and collecting specimens. In this study, children with severe pneumonia met the clinical case definition of severe pneumonia but had negative blood cultures; IPD cases met any one of the 4 clinical case definitions and had a positive blood and/or CSF culture for *S. pneumoniae*.

### Study design

To achieve the study objectives, we used two different study design approaches: (1) a before-after study in Mirzapur based on community surveillance for suspected pneumococcal disease and hospital case detection of blood culture-proven IPD at population level to compare: a) overall, vaccine type (VT) and non-vaccine type (NVT) IPD incidence, and b) incidence of severe pneumonia, and (2) an indirect cohort study design (using four sites, including the Mirzapur site) to assess the effectiveness of the PCV-10 against VTs, with IPD cases identified at KWMCH in Mirzapur and three other WHO - supported Invasive Bacterial Vaccine-Preventable Disease Surveillance Network sentinel site hospitals, namely, Bangladesh Shishu Hospital and Institute (formerly named as Dhaka Shishu Hospital), Dr. MR Khan Shishu Shasthya Foundation Hospital and Institute of Child Health, and Chittagong Ma O Shishu Hospital.

### Before-after comparison of incidence of IPD among 2- <24-month-old children

For calculation of IPD incidence in Mirzapur during the pre- and post-PCV10 introduction, IPD cases among children aged 2 – <24 months who presented to KWMCH were identified using standard laboratory methods for culture and identification of pneumococci from blood or CSF (14). The number of total IPD cases identified among 2 - <24-month-old children was used as the numerator and child-years observed was used as denominator for estimation of overall crude incidence. Incidence of VT and NVT IPD cases were calculated similarly using subsets of IPD cases. To measure changes in incidence rates, we compared incidence for four post-vaccine years (April 2015 - March 2019) with that of three pre-vaccine years (April 2012 - March 2015).

### Comparison of incidence of severe pneumonia between pre-and post-vaccine periods among children 2 - <24 months old

Like IPD incidence, we measured the crude incidence of severe pneumonia identified clinically among all children aged 2- <24 months who came to KWMCH from the study area (self-referred and VHW-referred). The number of severe pneumonia cases in this population was considered as the numerator and child-years observed as the denominator. The crude incidence was adjusted based on failure of VHW-referred cases to come to the study hospital out-patient department.

### Indirect Cohort Analysis

To calculate vaccine effectiveness, vaccination status for children with VT-IPD at all four study hospitals was compared with vaccination status of children with NVT-IPD; the method assumes that vaccination does not impact the risk for non-vaccine-type disease among vaccinated individuals. A recent analysis found that results using the indirect cohort method were comparable to that of a standard case-control study (15), which is complex and difficult to implement. In this analysis, VT-IPD (PCV10) was defined as cases of IPD in whom PCV10 serotypes and 6A (known to be cross-protected by PCV10) were identified. All other serotypes were considered NVT. All IPD cases with known serotypes collected from the four WHO sentinel hospitals from April 2015 through March 2019 were used for this analysis. Based on the card-verified immunization status, children with IPD were categorized into two groups: vaccinated and unvaccinated. The unvaccinated cohort included verbally confirmed non-vaccinated children and 2 - <24-month-old children who were not age-eligible for PCV at the time of the initial pneumococcal vaccination program (21^st^ March 2015). As Bangladesh’s EPI did not have catch-up campaigns and only children aged 6 weeks were eligible for vaccine introduction, this strategy led to an increase of vaccine eligible children gradually with time. To identify vaccine eligible children, we back calculated the birthdate of children and considered those born on 7^th^ February 2015 or later as eligible to receive the 1^st^ dose of pneumococcal vaccine at 6 weeks of age. Children who sought medical care two weeks or more after receiving an initial dose were considered vaccinated. Any child who came to the hospital with IPD within <14 days of receiving the 1^st^ dose of PCV10 vaccine was considered non-vaccinated. Children who received two or three doses of PCV-10 were considered individually for calculating vaccine impact. We calculated the odds ratio of receipt of PCV10 dose among children who had vaccine-type (PCV10) IPD versus non-vaccine-type IPD and used logistic regression to estimate vaccine effectiveness using the formula: *Vaccine effectiveness = (1-odds ratio for PCV10 vaccination) x 100%*.

### Laboratory methods

Blood specimens were cultured using BACTEC bottles. Beep positive and/or chocolate-colored blood bottles were screened by culture technique on blood and chocolate agar plates. CSF specimens were plated directly on blood and chocolate agar plates. In addition, all CSF specimens underwent testing for pneumococcus using an immunochromatographic method (BinaxNow [14]). Pneumococcal isolates were serotyped using capsular swelling method (quellung reaction). All BinaxNow positive specimens were tested for the lytA gene of pneumococcus and serotyped using previously published seven multiplex (triplex) and three in-house multiplex (triplex) RT-PCR assays covering a total of fifty serotypes (1, 2, 3, 4, 5, 6A/6B/6C/6D, 6C/6D, 7A/7F, 7B/C, 8, 9V/9A, 10A/F, 11A/D, 12F/12A/12B/44/46, 13, 14, 15A/B/C/F, 15A/15F, 16F, 18C/18A/18B/18F, 19A, 19F, 20, 22F/22A, 23A, 23F, 33F/33A/37, 34, 35B, 45 and 48) (16).

All laboratory methods were conducted at the respective four WHO sentinel site laboratories except the RT-PCR and capsular swelling, which were done at Bangladesh Shishu Hospital and Institute.

### Statistical analysis

Statistical analyses were conducted using STATA 14.0 (Stata Corp LP, College Station, TX). Results were reported as frequencies and percentages for categorical variables. We calculated incidence rates (per 100,000 child years of observation) and incidence rate ratios (IRR) using Poisson regression models for VT, NVT, overall IPD and physician confirmed severe pneumonia cases. The crude incidence calculations were adjusted for rate of compliance with VHW’s referral from the community to the hospital. The corresponding confidence intervals for each IRR and adjusted IRR were also calculated including their respective p-values. The level of significance was set at 5%, and p-values <0.05 were considered statistically significant.

### Ethical considerations

The study protocol was reviewed and approved by an independent Ethical Review Board based at Bangladesh Institute of Child Health, Dhaka, Bangladesh. Informed written consent was obtained by VHWs from the parents of the children before enrollment in community surveillance in Mirzapur. Informed consent was also obtained for enrollment in hospital surveillance at all four study hospitals, including collection of clinical data and specimens.

## Results

### Comparison of incidence of IPD pre- and post PCV10 introduction in Mirzapur

During the seven-year study period, we enrolled 20,187 children, including 3,395 in the pre-vaccine period and 16,792 in the post-vaccine period, who were monitored for illness suggestive of pneumonia or severe pneumonia as well as suspected sepsis and meningitis (Table 1). In total, these children contributed 34,608 (pre-vaccine, n=9,153 and post-vaccine, n=25,455) child-years of observation. In the baseline period, 500,295 visits were made by VHWs. Among assessed children, VHWs referred 2,589 children to KWMCH based on the referral criteria (Table 1); of these, 1327 (51%) complied and sought care at KWMCH. In the post-vaccine period, 12,830 children were referred by VHWs and 9,437 (73.5%) visited KWMCH (Supplemental Table 1).

During the pre-vaccine period, 2,617 children (1327 referred by VHWs and 1290 who sought care on their own) met the eligibility criteria at KWMCH for suspected pneumococcal disease (pneumonia, severe pneumonia, meningitis, or sepsis). Among these, 1,857 (71.0%) children were enrolled and had specimens and data collected. Five (0.27%; 5/1,857) blood samples were positive for pneumococci, four of which were VTs (80.0%). During the post-vaccine period, study physicians identified 8,342 children with suspected pneumococcal diseases. Of these, 89.8% (n=7487) were enrolled and subjected to blood culture. A total of nineteen pneumococci (0.25%; 19/7487) were detected, including nine vaccine serotypes (47.3%) and ten non-vaccine serotypes (52.7%; Figure 2).

**Figure 2:**
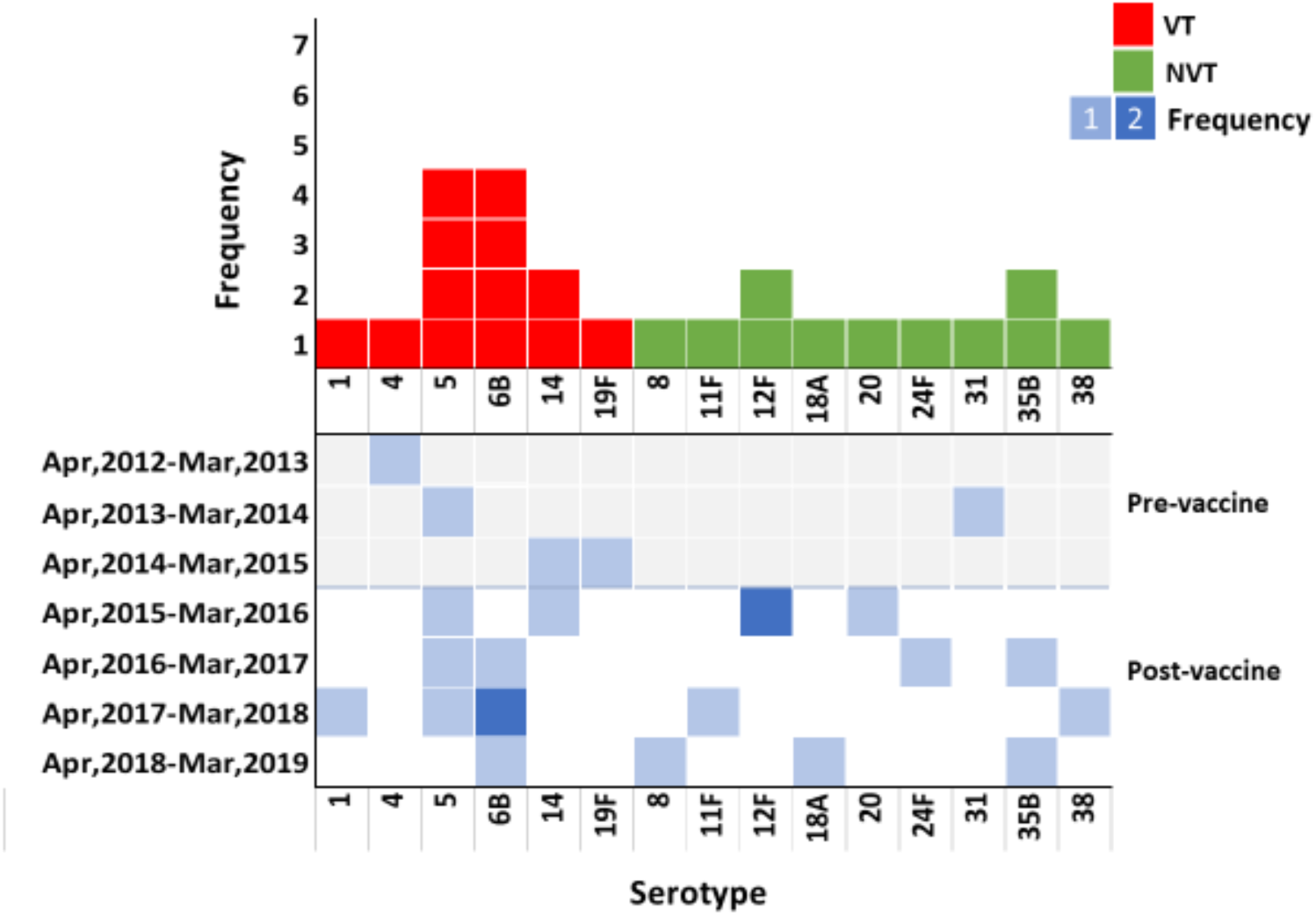
Serotype distribution of invasive pneumococcal cases with epidemiological year.

During the pre-PCV period, the crude incidence of invasive pneumococcal disease among children 2- <24 months of age in Mirzapur was 54.6 (95% CI: 6.8-102.5) cases per 100,000 child-years observed. VT-IPD incidence was 43.7 (95% CI: 0.9-86.5) and NVT-IPD was 10.9 (95%CI: 10.5-32.3) cases per 100,000 child-years. The average crude incidence of VT-IPD in the post-PCV period was 35.6 cases per 100,000 child-years observed (95% CI: -9.6-80.8). During the fourth-year post PCV introduction (April 2018 – March 2019), VT-IPD declined by 66% from the pre-vaccine period; VT cases reached 14.8 (95%CI: -14.2 to 43.9) and NVT cases reached 44.5 per 100,000 child-years (Figure 3A).

**Figure 3:**
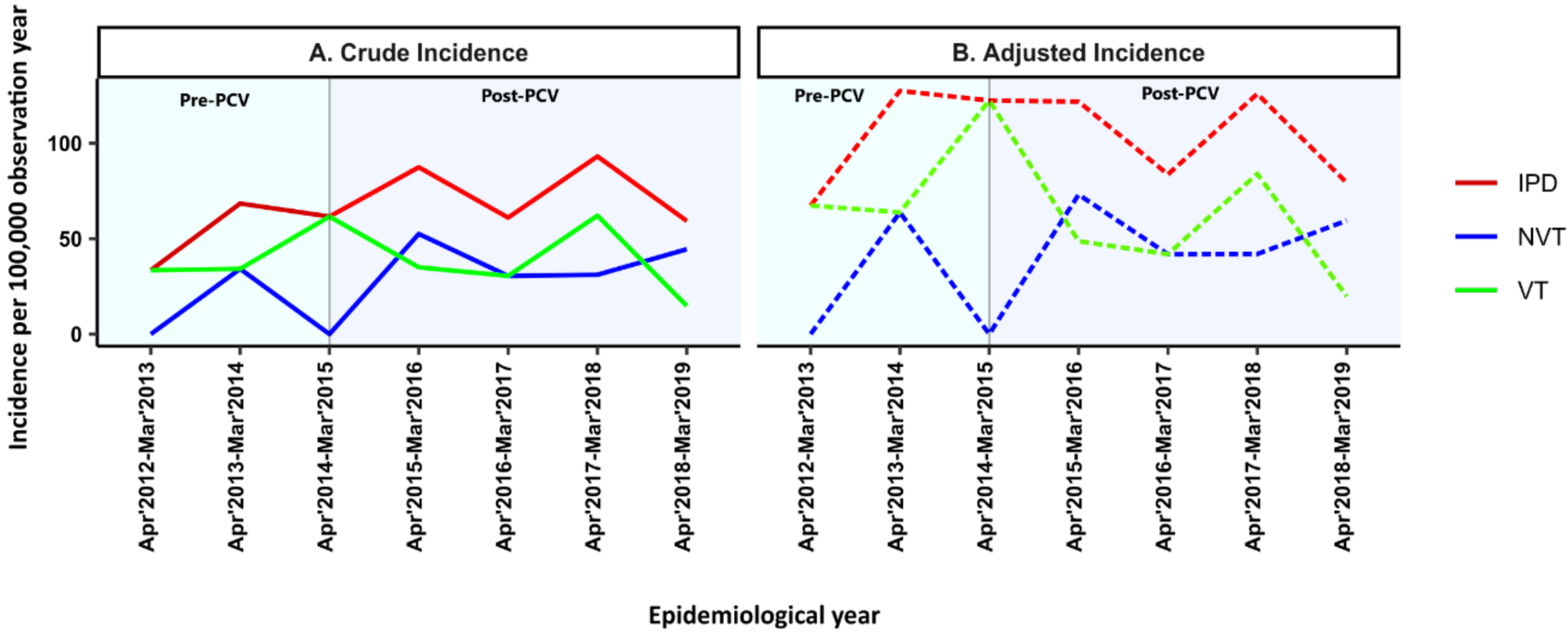
Changes in incidence for overall, vaccine type and non-vaccine type invasive pneumococcal disease between pre and post vaccine periods; A. Crude incidence. B. Adjusted incidence.

After adjusting for compliance with referrals, average incidence of IPD in the pre-PCV period was 106.1 (95% CI: 39.4-172.8) and post-PCV was 79.3 (95% CI: 12.1-146.6) cases per 100,000 child-years of observation (Figure 3B). Overall, IPD caused by any serotype declined 25% (p=0.58) among children under age 2 years. Comparing VT-IPD in the pre- and post-PCV10 periods, we observed a gradual decline of VT-IPD throughout the study period, with adjusted incidence reaching 19.8 cases in the final (4^th^) year from 85.7/100,000 child-years at baseline, a 77% decline (IRR 0.2; 95%CI: -0.04-1.45; p=0.068). During the 4^th^ year of the post-PCV period, incidence of NVT-IPD was 59.5 cases (95% CI: 1.3-117.7) per 100,00 child-years of observation, an increase from 20.4 cases in the pre-vaccine period (IRR: 2.9, 95% CI: 0.5-16.6; p=0.21). The increase in NVT-IPD was not driven by any specific serotype (Figure 2).

### Comparison of incidence of severe pneumonia pre-and post-PCV-10 introduction in Mirzapur

During the baseline period (April 2012 to March 2015), the average crude incidence of severe pneumonia cases was 50.1/1,000 (95% CI: 45.6-54.7) child-years observed. In the fourth year of the post-PCV period, incidence of severe pneumonia was lower by 33% (n= 33.8; 95%CI: 21%-44%, p-value <0.001) compared to baseline. After adjusting for compliance with referrals, the reduction of severe pneumonia was 54% (95% CI: 47%-59%, p-value <0.001), from 97.4 (95%CI: 91.0-103.8) cases at baseline to 45.2 cases (95%CI: 40.1-50.3)/1,000 child-years observed in year 4 (Figure 4).

**Figure 4:**
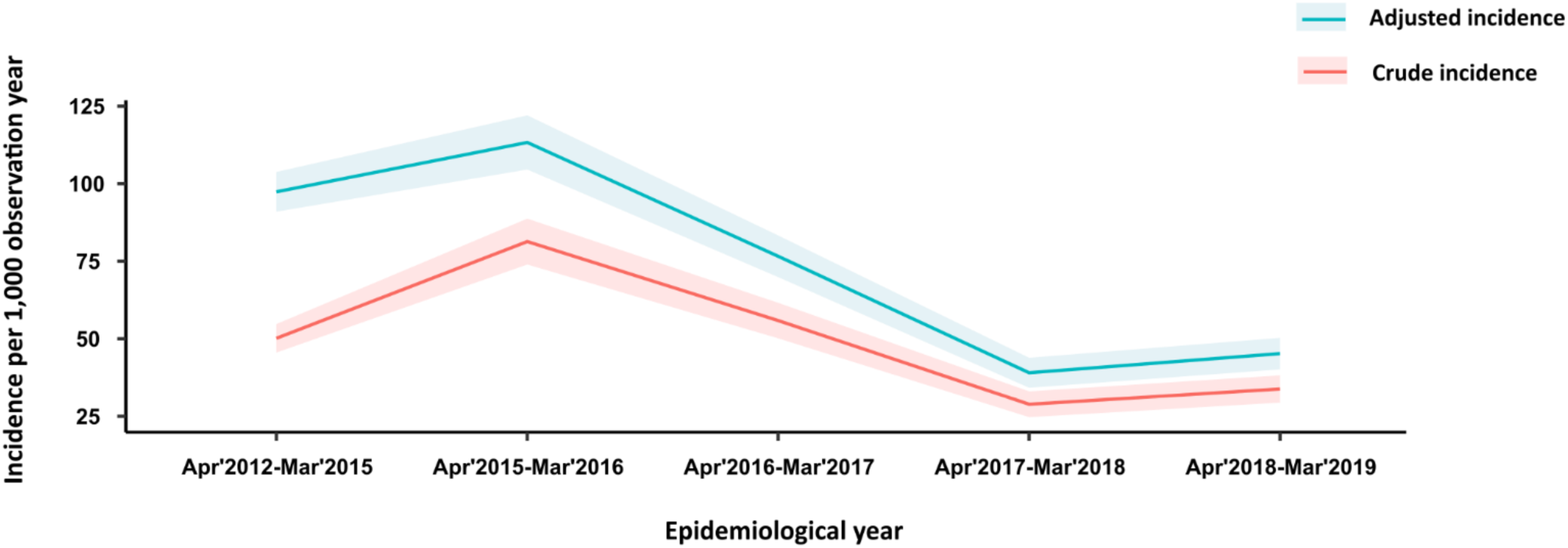
Crude and adjusted incidence of severe pneumonia cases seen at KWMCH among demographic surveillance area residents 2-<24 months of age. CI area showed in matched color shadow.

### PCV10 effectiveness against IPD in four sentinel sites

During the post-vaccine period (23^rd^ March 2015- 31^st^ March 2019), 171 IPD cases were detected from children evaluated at the four sentinel sites in Bangladesh (Supplementary figure-1). Among these children, 140 were eligible for PCV10 based on their age. Serotype information was available for 118 (84.2%, 118/140) cases, including 21 VT cases and 97 NVT cases. Among these 118 cases, 96 children with 14 VT disease and 82 with NVT disease were vaccinated and 22 with 7 cases of VT disease and 15 with NVT disease were unvaccinated. The median age of vaccinated children and unvaccinated children were 5 and 3 months, respectively. Effectiveness of the 1^st^ PCV10 dose was 37.0% (95% CI: -141.0% - 83.5%; p-value =0.49). Two or three doses gave protection of about 63.1% (95% CI: 3.3% – 85.9%; p-value =0.0411) against VT IPD.

## Discussion

IPD is a major cause of illness in young children in Bangladesh, with 88% (2007-2013) of cases in children aged <2 years (11). Since this cohort is the main target of the conjugate vaccine, estimating the effectiveness of pneumococcal vaccine in this vulnerable age group (under 2 years) is crucial. In addition, understanding the true burden of pneumococcal diseases in Bangladesh is difficult because most children receive antibiotics before cultures are taken and blood cultures have low sensitivity for determining pneumonia etiology, even among children who are not previously treated. Therefore, we measured the impact of PCV10 by examining trends in both culture-confirmed pneumococcal disease and physician confirmed severe pneumonia cases.

Our population-based study revealed that PCV10 introduction was associated with a 77% reduction in vaccine type (PCV10+6A) IPD and a 25% reduction in overall IPD during the first 4 years of the program, although these reductions were not statistically significant. The decrease in VT disease observed in this study is concordant with findings in other countries. For example, within two years of PCV7 introduction in the United States, VT and VT related serotypes declined by 78% (6). Similarly, South Africa achieved an 85% reduction rate among HIV uninfected children under 2 years of age within three years of introducing PCV (PCV7 and PCV13) (8). On the other hand, there have been variations in reduction of overall IPD depending on the diversity of serotypes. For instance, among children <2 years of age, a 69% decline of overall IPD was observed in both the United States and South Africa following two and four years of vaccine implementation, respectively (6, 8). In contrast, in Brazil, the rate of IPD declined 44% among children 2- <24 months old three years following PCV10 induction (17). In Bangladesh, the reduction of overall IPD was lower (25%) compared to what was measured in some settings [8-10] and like others. This is most likely due to the large diversity of serotypes causing IPD in Bangladesh (51 different invasive serotypes) along with the persistent >10-year dominance of serotype 2 that we documented (11). Catch-up programs with vaccination of children up to 2 years of age in the US and up to 24 months in the United Kingdom (UK) may have resulted in faster drops in VT cases than what we observed (6, 7, 9).

Our study demonstrated a statistically significant reduction (54%, p<0.001) of hospitalization with severe pneumonia for four years following vaccine introduction in this rural population. Although WHO proposes use of chest radiograph as a benchmark for evaluating the efficacy of pneumococcal vaccine (18), this also leads to underestimation of the impact of pneumococcal vaccine on the pneumonia burden by up to 63% (19). Similarly, a study of PCV10 vaccine effectiveness against radiologic pneumonia among vaccine eligible children in rural Bangladesh found 21.4% effectiveness among children 3-11 months of age who received 3 doses of PCV10 (20), lower than the impact we measured but similar to findings from a clinical trial evaluating PCV10 efficacy against radiological confirmed pneumonia (21). This low effectiveness is possibly due to high specificity that missed pneumococcal pneumonia cases (19) and serotype replacement. During this post PCV period, we found an increase of NVT-IPD incidence (IRR: 2.9; p-value= 0.21) compared to pre-PCV, which may have occurred in radiologic pneumonia cases as well. A possible reason of higher effectiveness in our study could be our selection of only severe pneumonia cases.

Consistent with other countries using PCV10, our indirect cohort analysis from multiple contemporary surveillance data showed that PCV10 provides significant protection (63.1%) with two or more doses of vaccine for under 2-year-old children (15, 22-24). A case-control study from Pakistan showed effectiveness of PCV10 to be 81.9% against vaccine-type IPD among completely vaccinated children <5 years (22). An indirect cohort analysis study in Brazil reported 72.8% effectiveness against VT among children <5 years who had received one or more doses (15). A serotype-specific effectiveness study from the UK revealed that PCV7 gave less protection (49%) against 6B compared to the other six serotypes (25, 26). Similar results were seen against 6B in the USA, where a PCV manufactured by Pfizer was used (27).

A strength of our study is the multiple approaches we were able to use to evaluate PCV10 benefits among children in Bangladesh. We not only measured PCV10 impact on the burden of two types of pneumococcal disease – IPD and severe pneumonia – using a population-based system, but we also assessed effectiveness in vaccinated children using an indirect cohort method in four hospitals with established hospital surveillance for IPD.

Multiple limitations should be considered. Overall, impact of PCV in preventing IPD was not significant, due to the small number of IPD cases during the study period. This low number of isolates also hindered our ability to measure the impact of vaccine on possible antimicrobial resistant strains and emergence of NVT in this population. We missed very severe pneumonia with/without hypoxemia cases, as the study was in our DSS area, where we have active surveillance and weekly home visits for wellbeing of under-five children, thus resulting in early case detection. Also, VHWs, while trained with attending physicians, might miss some of the suspected cases of pneumonia, sepsis, and meningitis. This could result in reduction in estimation of efficacy of the vaccine.

Despite these limitations, this study demonstrates that PCV10 is already reducing the morbidity and possible mortality attributed to pneumococcal infection in a high-risk rural population, including reducing a substantial number of hospitalizations with severe pneumonia. However, given the diversity of serotypes causing pneumococcal disease in Bangladesh, ongoing monitoring is required to capture emergence of NVT which could reduce vaccine impact in the coming years. Continued surveillance will also help policy makers to make need-based decisions about future formulations of pneumococcal vaccines. Finally, our findings should encourage the introduction of lifesaving PCVs in other countries where these vaccines have not yet been introduced.

## Data Availability

All data produced in the present study are available upon reasonable request to the authors.

## Author Contribution

## Acknowledgement

## Author’s declaration

The authors declare that they have no known competing financial interests or personal relationships that could have appeared to influence the work reported in this paper

---

**Supplementary Figure 1:**
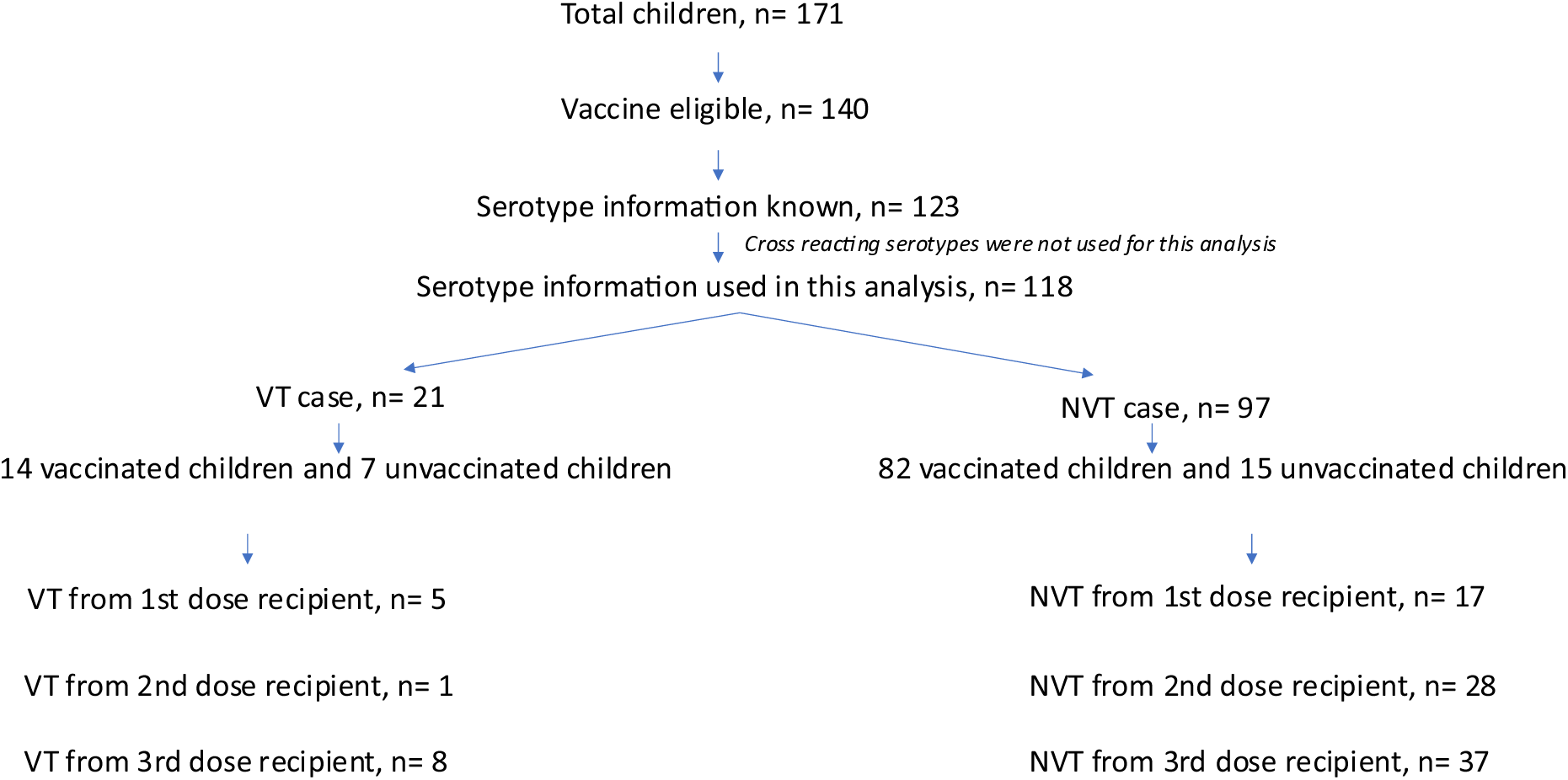
Vaccination status of all IPD cases.

**Supplementary Table 1:** Summary data of population and hospital based surveillance.

| Type of surveillance | Category | Pre-vaccine |  |  | Post-vaccine |  |  |  |
| --- | --- | --- | --- | --- | --- | --- | --- | --- |
|  |  | Apr,2012-Mar,2013 | Apr,2013-Mar,2014 | Apr,2014-Mar,2015 | Apr,2015-Mar,2016 | Apr,2016-Mar,2017 | Apr,2017-Mar,2018 | Apr,2018-Mar,2019 |
| Community based surveillance | No of Enrolled children | 1083 | 1039 | 1273 | 2542 | 3134 | 4046 | 7070 |
|  | Child years of observation | 2985 | 2922 | 3246 | 5717 | 6552 | 6441 | 6745 |
|  | No of follow-up by VHW | 212375 | 161507 | 126413 | 247460 | 386735 | 510815 | 646508 |
|  | Episodes assessed by VHW | 10919 | 11060 | 11718 | 20136 | 27671 | 31738 | 30837 |
|  | Children referred by VHW | 927 | 883 | 779 | 2311 | 3085 | 3762 | 3672 |
|  | Children visited to KWMCH | 461 | 474 | 392 | 1659 | 2250 | 2783 | 2745 |
|  | Compliance rate, % | 50% | 54% | 50% | 72% | 73% | 74% | 75% |
| Hospital based surveillance | No of eligible children (self and VHW referred) | 1040 | 880 | 697 | 1966 | 2431 | 2046 | 1899 |
|  | Diagnosed with pneumonia | 273 | 218 | 187 | 776 | 1,127 | 555 | 625 |
|  | 2-5 months (%) | 77 (28%) | 53 (24%) | 39 (21%) | 237 (31%) | 291 (26%) | 185 (33%) | 162 (26%) |
|  | 6-11 months (%) | 86 (31%) | 75 (34%) | 59 (32%) | 280 (36%) | 401 (36%) | 195 (35%) | 241 (39%) |
|  | 12-23 months (%) | 110 (40%) | 90 (41%) | 89 (48%) | 259 (33%) | 435 (39%) | 175 (32%) | 222 (36%) |
|  | Diagnosed with severe pneumonia | 131 | 192 | 136 | 465 | 366 | 186 | 228 |
|  | 2-5 months (%) | 59 (45%) | 86 (45%) | 49 (36%) | 202 (43%) | 112 (31%) | 72 (39%) | 78 (34%) |
|  | 6-11 months (%) | 40 (31%) | 50 (26%) | 40 (30%) | 136 (29%) | 134 (37%) | 60 (32%) | 68 (30%) |
|  | 12-23 months (%) | 32 (25%) | 56 (29%) | 47 (35%) | 127 (27%) | 120 (33%) | 54 (29%) | 82 (36%) |
|  | **Diagnosed with meningitis** | 56 | 68 | 99 | 86 | 89 | 84 | 54 |
|  | **2-5 months (%)** | 3 (5%) | 6 (8%) | 19 (19%) | 13 (15%) | 8 (9%) | 10 (12%) | 2 (4%) |
|  | **6-11 months (%)** | 14 (25%) | 20 (30%) | 32 (32%) | 27 (31%) | 25 (29%) | 24 (29%) | 22 (41%) |
|  | **12-23 months (%)** | 39 (70%) | 42 (62%) | 48 (49%) | 46 (53%) | 56 (63%) | 50 (60%) | 30 (56%) |
|  | **No of blood culture from eligible children** | 746 | 654 | 451 | 1630 | 2231 | 1882 | 1744 |
|  | **No of pneumococcal isolates** | 1 | 2 | 2 | 5 | 4 | 6 | 4 |
|  | **No of VT** | 1 | 1 | 2 | 2 | 2 | 4 | 1 |
|  | **No of NVT** | 0 | 1 | 0 | 3 | 2 | 2 | 3 |

## Reference

1. Organization WH. Pneumococcal disease 2020 [Available from: https://www.who.int/immunization/diseases/pneumococcal/en/.

2. O’Brien KL, Wolfson LJ, Watt JP, Henkle E, Deloria-Knoll M, McCall N, et al. Burden of disease caused by Streptococcus pneumoniae in children younger than 5 years: global estimates. Lancet. 2009;374(9693):893-902.

3. Wahl B, O’Brien KL, Greenbaum A, Majumder A, Liu L, Chu Y, et al. Burden of Streptococcus pneumoniae and Haemophilus influenzae type b disease in children in the era of conjugate vaccines: global, regional, and national estimates for 2000-15. Lancet Glob Health. 2018;6(7):e744–e57.

4. Collaborators GBDLRI. Estimates of the global, regional, and national morbidity, mortality, and aetiologies of lower respiratory infections in 195 countries, 1990-2016: a systematic analysis for the Global Burden of Disease Study 2016. Lancet Infect Dis. 2018;18(11):1191-210.

5. Organization WH. Pneumococcal conjugate vaccine for childhood immunization--WHO position paper. Wkly Epidemiol Rec. 2007;82(12):93–104.

6. Whitney CG, Farley MM, Hadler J, Harrison LH, Bennett NM, Lynfield R, et al. Decline in invasive pneumococcal disease after the introduction of protein-polysaccharide conjugate vaccine. N Engl J Med. 2003;348(18):1737–46.

7. Waight PA, Andrews NJ, Ladhani SN, Sheppard CL, Slack MP, Miller E. Effect of the 13-valent pneumococcal conjugate vaccine on invasive pneumococcal disease in England and Wales 4 years after its introduction: an observational cohort study. Lancet Infect Dis. 2015;15(5):535–43.

8. von Gottberg A, de Gouveia L, Tempia S, Quan V, Meiring S, von Mollendorf C, et al. Effects of vaccination on invasive pneumococcal disease in South Africa. N Engl J Med. 2014;371(20):1889–99.

9. Ladhani SN, Collins S, Djennad A, Sheppard CL, Borrow R, Fry NK, et al. Rapid increase in non-vaccine serotypes causing invasive pneumococcal disease in England and Wales, 2000-17: a prospective national observational cohort study. Lancet Infect Dis. 2018;18(4):441-51.

10. Pneumococcal conjugate vaccine for childhood immunization--WHO position paper. Wkly Epidemiol Rec. 2007;82(12):93-104.

11. Saha SK, Hossain B, Islam M, Hasanuzzaman M, Saha S, Hasan M, et al. Epidemiology of Invasive Pneumococcal Disease in Bangladeshi Children Before Introduction of Pneumococcal Conjugate Vaccine. Pediatr Infect Dis J. 2016;35(6):655–61.

12. Saha SK, Naheed A, El Arifeen S, Islam M, Al-Emran H, Amin R, et al. Surveillance for invasive Streptococcus pneumoniae disease among hospitalized children in Bangladesh: antimicrobial susceptibility and serotype distribution. Clin Infect Dis. 2009;48 Suppl 2:S75–81.

13. Arifeen SE, Saha SK, Rahman S, Rahman KM, Rahman SM, Bari S, et al. Invasive pneumococcal disease among children in rural Bangladesh: results from a population-based surveillance. Clin Infect Dis. 2009;48 Suppl 2:S103–13.

14. Cheesbrough M. District Laboratory Practice in Tropical Countries. 2nd ed. United Kingdom: Cambridge University Press; 2006.

15. Verani JR, Domingues CM, de Moraes JC, Brazilian Pneumococcal Conjugate Vaccine Effectiveness Study G. Indirect cohort analysis of 10-valent pneumococcal conjugate vaccine effectiveness against vaccine-type and vaccine-related invasive pneumococcal disease. Vaccine. 2015;33(46):6145-8.

16. Pimenta FC, Roundtree A, Soysal A, Bakir M, du Plessis M, Wolter N, et al. Sequential triplex real-time PCR assay for detecting 21 pneumococcal capsular serotypes that account for a high global disease burden. J Clin Microbiol. 2013;51(2):647-52.

17. Andrade AL, Minamisava R, Policena G, Cristo EB, Domingues CM, de Cunto Brandileone MC, et al. Evaluating the impact of PCV-10 on invasive pneumococcal disease in Brazil: A time-series analysis. Hum Vaccin Immunother. 2016;12(2):285–92.

18. Cherian T, Mulholland EK, Carlin JB, Ostensen H, Amin R, de Campo M, et al. Standardized interpretation of paediatric chest radiographs for the diagnosis of pneumonia in epidemiological studies. Bull World Health Organ. 2005;83(5):353–9.

19. Madhi SA, Klugman KP. World Health Organisation definition of "radiologically-confirmed pneumonia" may under-estimate the true public health value of conjugate pneumococcal vaccines. Vaccine. 2007;25(13):2413–9.

20. McCollum ED, Ahmed S, Roy AD, Chowdhury NH, Schuh HB, Rizvi SJR, et al. Effectiveness of the 10-valent pneumococcal conjugate vaccine against radiographic pneumonia among children in rural Bangladesh: A case-control study. Vaccine. 2020;38(42):6508–16.

21. Tregnaghi MW, Saez-Llorens X, Lopez P, Abate H, Smith E, Posleman A, et al. Efficacy of pneumococcal nontypable Haemophilus influenzae protein D conjugate vaccine (PHiD-CV) in young Latin American children: A double-blind randomized controlled trial. PLoS Med. 2014;11(6):e1001657.

22. Riaz A, Mohiuddin S, Husain S, Yousafzai MT, Sajid M, Kabir F, et al. Effectiveness of 10-valent pneumococcal conjugate vaccine against vaccine-type invasive pneumococcal disease in Pakistan. Int J Infect Dis. 2019;80:28–33.

23. Domingues CM, Verani JR, Montenegro Renoiner EI, de Cunto Brandileone MC, Flannery B, de Oliveira LH, et al. Effectiveness of ten-valent pneumococcal conjugate vaccine against invasive pneumococcal disease in Brazil: a matched case-control study. Lancet Respir Med. 2014;2(6):464–71.

24. Peckeu L, van der Ende A, de Melker HE, Sanders EAM, Knol MJ. Impact and effectiveness of the 10-valent pneumococcal conjugate vaccine on invasive pneumococcal disease among children under 5 years of age in the Netherlands. Vaccine. 2021;39(2):431–7.

25. Andrews N, Waight PA, Borrow R, Ladhani S, George RC, Slack MP, et al. Using the indirect cohort design to estimate the effectiveness of the seven valent pneumococcal conjugate vaccine in England and Wales. PLoS One. 2011;6(12):e28435.

26. Pichon B, Ladhani SN, Slack MP, Segonds-Pichon A, Andrews NJ, Waight PA, et al. Changes in molecular epidemiology of streptococcus pneumoniae causing meningitis following introduction of pneumococcal conjugate vaccination in England and Wales. J Clin Microbiol. 2013;51(3):820–7.

27. Park SY, Van Beneden CA, Pilishvili T, Martin M, Facklam RR, Whitney CG, et al. Invasive pneumococcal infections among vaccinated children in the United States. J Pediatr. 2010;156(3):478–83 e2.

